# Prediction of Stature using Percutaneous long bones of the Upper and Lower limbs among Asante and Ewe Ethnic Groups in Ghana

**DOI:** 10.1101/2024.12.14.24319028

**Authors:** Daniel Kobina Okwan, Chrissie Stansie Abaidoo, Pet-Paul Wepeba, Juliet Robertson, Samuel Kwadwo Peprah Bempah, Priscilla Obeng, Ethel Akua Achiaa Domfeh, Sarah Owusu Afriyie, Thomas Kwaku Asante

## Abstract

**Introduction:** Sophisticated technological advancements for identification of people are readily available in developed countries. Meanwhile, relatively less expensive algorithms in physical anthropometry could be employed for such identification purposes. Although such evaluations have been done in some countries, due to interpopulation variations, such relations should be ethnic-and sex-specific.

**Aim:** Therefore, the present study sought to assess the relationship between long bones of upper and lower limbs among two ethnic groups in Ghana for stature estimation.

**Methodology:** Using a purposive non-random sampling technique, participants made up of 140 Asantes and 102 Ewes aged 20 to 25 years were recruited after an ethical approval was obtained from the Committee on Human Research, Publication and Ethics, KNUST.

**Results:** For the same sex, there was no statistically significant difference between Asantes and Ewes with respect to height. The most useful parameter for stature estimation among the Asante males was left fibular and tibial lengths with that of the females being left ulnar, fibular and radial lengths. However, for the Ewe males, the most significant parameters for height estimation were right fibular and humeral lengths with the Ewe females being right tibial and humeral lengths.

**Conclusion:** Findings of the study are useful for the identification of humans with dismembered body parts involved in various disasters such as automobile accidents. The database and formulae derived would be useful for stature estimation needed in biological profiling and other assessments of bedridden patients.

## Introduction

It has been reported that human identification of unknown remains is commonly done using long bones. This is because, employing various anthropometric algorithms, stature of the deceased could be estimated from the remains of the skeletal parts of the body [1]. The alternative means of doing this is to employ percutaneous measurement of bones [2].

Moreover, anthropometric means of predicting stature and sex is more practical and cost-effective compared to DNA analysis [3]. Apart from sex, age and race, stature plays a significant role during the identification of people [4]. Particularly for people with disproportionate growth problem, myopathy, skeletal dysplasia, or stature loss during spinal surgeries, predictions of stature using measurements of body parts are often helpful [5]. Furthermore, regression models developed for height estimation utilizing long bones frequently have significantly lower standard errors of estimates. Long bones are therefore more accurate at measuring stature [6,7].

According to Dumitri *et al.* [8], external variables that can alter bone formation include diet, exercise and psychosocial factors. The nature of occupation or task one is involved in, lifestyle particularly due to the influence of civilisation may be impactful on body parts, including the limbs [1].

In predicaments that result in mutilated body parts, medicolegal experts are challenged to help identify the victims [9,10]. In fact, the events of natural calamities continue to increase every now and then with mutilated body parts frequently encountered [11]. In Ghana, reports show that there is increased natural disasters and road traffic accidents resulting in multiple casualties [12]. The identification is usually resolved using tested formulae which are specific to one population and therefore cannot be applied to another population since variations exist owing to diet, environment and lifestyle [13].

With the increasing number of medicolegal cases, comprehensive knowledge of the modern human populations in the Ghanaian society has become urgent so that correct interpretation of unknown persons from contemporary forensic applications can be provided. Therefore, there is the utmost need to obtain ethnic-specific identification formulae for the assessment of the relationship between long bones of the limbs and height. Thus, the current study sought to analyze and bring out the association between percutaneous lengths of the limbs among Asante and Ewe ethnic groups in Ghana using percutaneous approach for stature estimation.

## Materials and Methods

### Study Design and Ethical Issues

This study employed a cross-sectional design to recruit participants aged 20 – 25 years. Ethical approval was sought from Committee on Human Research, Publication and Ethics, School of Medical Sciences, Kwame Nkrumah University of Science and Technology (KNUST), Kumasi, Ghana (Reference Number: CHRPE/AP/129/20). All procedures performed on the participants were done in accordance with the ethical standards outlined in the 1964 and subsequent modifications of the Declaration of Helsinki. Informed consent was as well sought from each study participant before relevant data were taken confidentially. A non-probability purposive sampling technique was employed to recruit pure (both parents and grandparents from the ethnic group of interest) Ewe and Asante participants. The study was made up of 140 Asantes [71(50.7%) males and 69(49.3%) females] and 102 Ewes [61(59.8%) males and 41(40.2%) females].

### Inclusion Criteria

Only consented participants from the aforementioned ethnic groups aged 20 - 25 years were recruited for the present study.

### Exclusion Criteria

Individuals with old or current fracture(s) of the upper or lower limb or other orthopaedic deformities (kyphosis, lordosis, scoliosis) that could affect the measurements of the long bones or stature were excluded. Also, individuals with metabolic or developmental disorders (dwarfism, gigantism) than could potentially affect the maximum dimensions of the bones or stature were excluded from the present study.

### Measurements Taken

For both sexes and ethnic groups, with the exception of the stature, each remaining parameter was taken bilaterally following the protocols suggested in the International Standards for Anthropometric Assessment published by the International Society for the Advancement of Kinanthropometry (ISAK).

**Humeral length:** Percutaneous distance from the lateral end of the clavicle to the lateral epicondyle of the humerus. This was done with the elbow bent at an angle of 90° and participants being in siting position and the arm fully adducted at the shoulder joint.

**Ulnar length:** It was measured as the distance from the tip of the olecranon process of the ulna to the head of the ulna. In other words, it is the displacement of the styloid process from the tip of the olecranon process with the elbow fully flexed (90°) and palm spread over the opposite shoulder.

**Radial length**: It was measured as the displacement from the radial head to the radial styloid process.

**Femoral length**: It was measured as the displacement from the greater trochanter to the lateral epicondyle of the femur.

**Tibial length:** It was measured as the distance from the medial-most superficial point on the upper border of the medial condyle of the tibia to the tip of the medial malleolus (Spherion) of the tibia. The angle between flexor surface of the leg and the thigh was maintained at 90°. This was achieved by asking each participant to sit (as it is easier to access the tibia in this position) facing the observer with ankle resting on the knee, so that the medial aspect of the tibia faced upwards.

**Fibular length:** It was measured as the distance from the fibular head to the lateral malleolus. Each participant was asked to sit in order to make the landmarks easily accessible and palpable.

**Stature of the participants**: It was taken using a stadiometer as the displacement from the vertex of the head to the floor with participants being barefooted, feet close together, body erect and head positioned in the Frankfurt horizontal plane in accordance with the International Biological Programme [14]. The shoulders of each participant were relaxed, the back was made straight, upper surface of the thighs made horizontal, the feet supported and the back of the knee joint cleared of any obstacle and then the vertex of the head contacted firmly with the stadiometer before the value was recorded.

### Precautions observed during measurements

To reduce both inter-observer and intra-observer rate of error and to ensure reproducibility, all measurements were taken twice by the same investigator. Long bone lengths and stature of all participants were taken at a specific time from 10.00 am to 2.00 pm. Most of the landmarks were highlighted on the skin using a marking pencil to increase accuracy.

### Definition of Key Concepts

**Percutaneous** refers to that which takes place through the skin or with the skin present.

**Anthropometry** refers to the measurement of some areas of the human body.

**Ethnicity** refers to the fact of belonging to a population group or subgroup consisting of persons who share a common cultural background.

**Stature** refers to the natural height of an individual while standing in an upright position.

### Statistical Analysis

Data obtained from study participants were entered into Microsoft Office Excel Spreadsheet 2019 and analyzed using IBM SPSS version 26.0. For descriptive statistics, data were expressed in means together with standard deviation. Independent samples t-test was employed to assess sexual dimorphism in the study participants based on parameters studied. Existence of bilateral asymmetry was assessed using paired samples t-test. Statistically significant level for differences or otherwise was pegged at p < 0.05 (95% confidence level). Pearson’s correlation coefficient (r) was used as the measure of strength and direction of the association between height and percutaneous bone lengths for both males and females of each ethnic group. Coefficient of determination (R^2^) together with its adjusted value (adjusted R^2^) was estimated to determine how much of the variance in the dependent variable could be explained by its relationship to the other variables. Using stepwise approach, linear regression equations (both simple and multivariate) were derived as predictive models for stature using percutaneous long bone lengths.

## Results

### Intrasex variation among Asantes and Ewes

#### Differences Between Anthropometric Parameters of Both Males and Females of Different Ethnic Groups

Using independent samples t-test, some variations between Asante males and Ewe males as well as Asante females and Ewe females were observed (Table 1).

**Table 1:** Comparison of Anthropometric Parameters of Both Males and Females of Different Ethnic Groups.

| | MALES [MEAN ± SD (cm)] | | | | FEMALES [MEAN ± SD (cm)] | | | |
| --- | --- | --- | --- | --- | --- | --- | --- | --- |
|  | Asantes<br>(N=71) | Ewes (N=61) | t | p | Asantes<br>(N=69) | Ewes (N=41) | t | p |
| RHL | 35.73 ± 0.98 | 35.63 ± 0.78 | 0.645 | 0.520 | 33.70 ± 1.49 | 33.14 ± 1.48 | 1.900 | 0.060 |
| RUL | 28.71 ± 1.24 | 29.36 ± 1.18 | -3.083 | 0.002 | 26.47 ± 1.55 | 27.05 ± 1.16 | -2.077 | 0.040 |
| RRL | 25.41 ± 1.59 | 26.27 ± 1.53 | -3.189 | 0.002 | 23.05 ± 1.46 | 24.07 ± 1.37 | -3.644 | 0.000 |
| RFL | 39.92 ± 1.70 | 39.52 ± 1.26 | 1.560 | 0.121 | 37.17 ± 1.47 | 38.32 ± 1.02 | -4.828 | 0.000 |
| RTL | 38.17 ± 1.62 | 38.41 ± 1.05 | -1.025 | 0.308 | 35.96 ± 1.64 | 37.28 ± 0.97 | -5.303 | 0.000 |
| RFibL | 36.88 ± 1.33 | 37.23 ± 0.97 | -1.688 | 0.094 | 34.85 ± 1.52 | 36.18 ± 0.99 | -5.518 | 0.000 |
| LHL | 35.85 ± 1.03 | 35.84 ± 0.81 | 0.087 | 0.931 | 33.77 ± 1.60 | 33.31 ± 1.49 | 1.490 | 0.139 |
| LUL | 28.73 ± 1.29 | 29.53 ± 1.11 | -3.843 | 0.000 | 26.56 ± 1.61 | 27.14 ± 1.24 | -1.985 | 0.050 |
| LRL | 25.53 ± 1.61 | 26.46 ± 1.46 | -3.444 | 0.001 | 23.04 ± 1.47 | 24.16 ± 1.53 | -3.815 | 0.000 |
| LFL | 39.66 ± 1.73 | 39.29 ± 1.33 | 1.334 | 0.185 | 36.93 ± 1.51 | 38.11 ± 1.04 | -4.424 | 0.000 |
| LTL | 37.85 ± 1.64 | 38.17 ± 1.18 | -1.260 | 0.210 | 35.87 ± 1.63 | 36.98 ± 0.97 | -4.479 | 0.000 |
| LFibL | 36.70 ± 1.51 | 37.04 ± 0.92 | -1.580 | 0.117 | 34.67 ± 1.56 | 35.98 ± 1.04 | -5.249 | 0.000 |

*RHL = Right Humeral Length; RUL = Right Ulnar Length; RRL = Right Radial Length; RFL = Right Femoral length; RTL = Right Tibial Length; RFibL = Right Fibular Length; LHL = Left Humeral Length; LUL = Left Ulnar Length; LRL = Left Radial Length; LFL = Left Femoral length; LTL = Left Tibial Length; LFibL = Left Fibular length; N = Number of Participants; SD = Standard Deviation; cm = centimetre; t = test statistic; p = Statistical significance level*

#### Bilateral differences between right and left measurements of Asante participants stratified by sex using paired samples t-test

Bilateral asymmetry of ethnic-and sex-specific parameters of upper and lower limbs are shown in Tables 2 and 3.

**Table 2:**
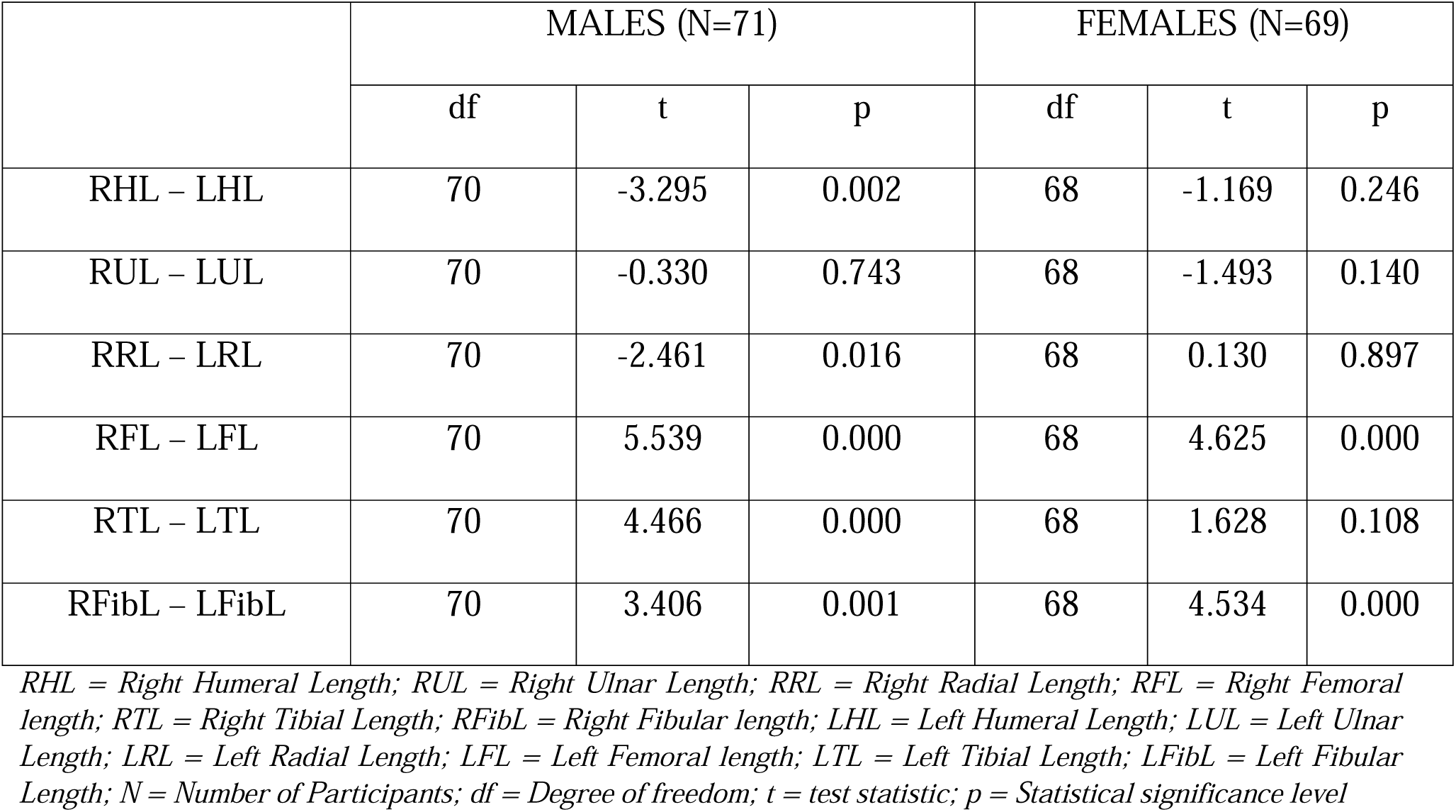
Comparison of Right and Left Anthropometric Parameters of Asantes Stratified by Sex.

|  | MALES (N=71) |  |  | FEMALES (N=69) |  |  |
| --- | --- | --- | --- | --- | --- | --- |
|  | df | t | p | df | t | p |
| RHL – LHL | 70 | -3.295 | 0.002 | 68 | -1.169 | 0.246 |
| RUL – LUL | 70 | -0.330 | 0.743 | 68 | -1.493 | 0.140 |
| RRL – LRL | 70 | -2.461 | 0.016 | 68 | 0.130 | 0.897 |
| RFL – LFL | 70 | 5.539 | 0.000 | 68 | 4.625 | 0.000 |
| RTL – LTL | 70 | 4.466 | 0.000 | 68 | 1.628 | 0.108 |
| RFibL – LFibL | 70 | 3.406 | 0.001 | 68 | 4.534 | 0.000 |

*RHL = Right Humeral Length; RUL = Right Ulnar Length; RRL = Right Radial Length; RFL = Right Femoral length; RTL = Right Tibial Length; RFibL = Right Fibular length; LHL = Left Humeral Length; LUL = Left Ulnar Length; LRL = Left Radial Length; LFL = Left Femoral length; LTL = Left Tibial Length; LFibL = Left Fibular Length; N = Number of Participants; df = Degree of freedom; t = test statistic; p = Statistical significance level*

**Table 3:**
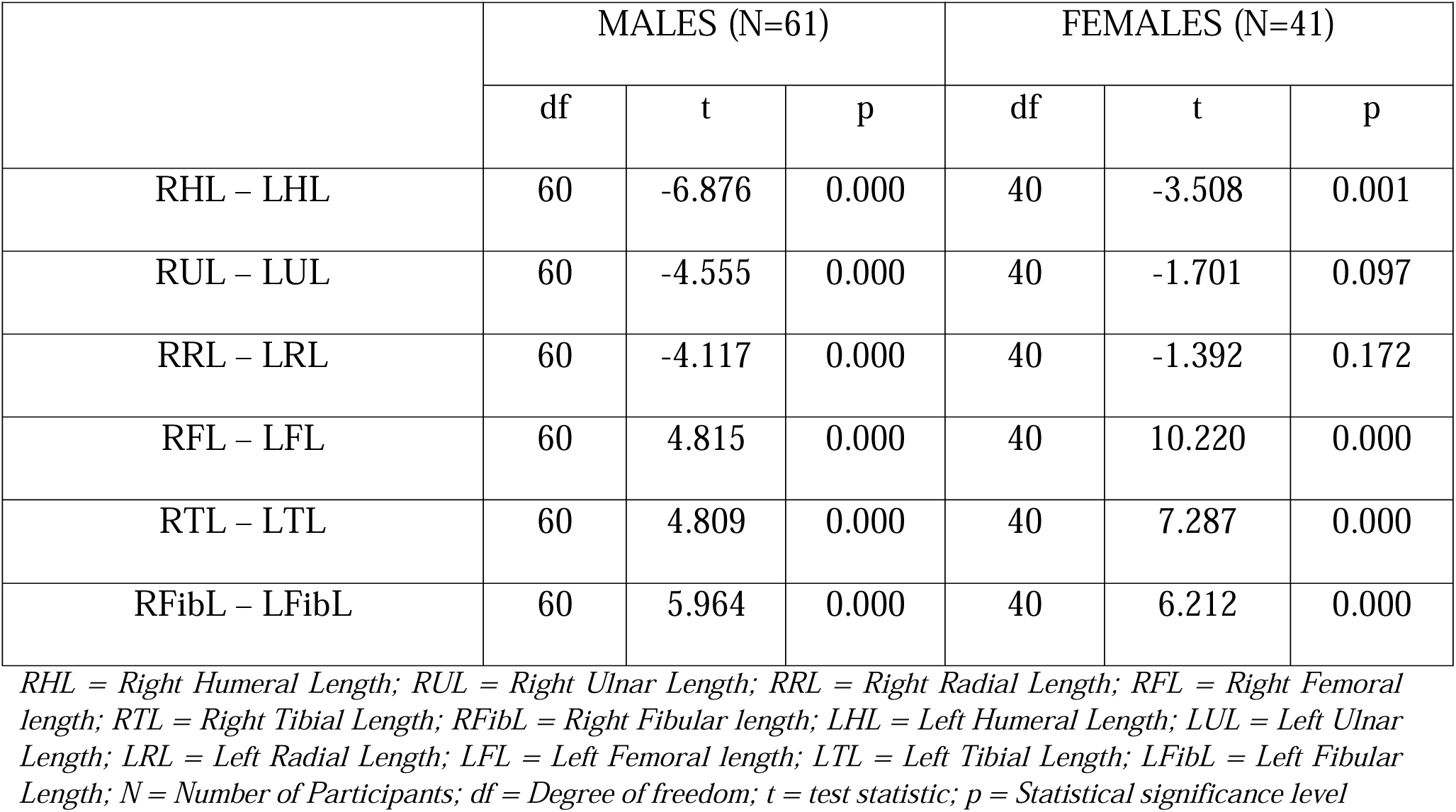
Comparison of Right and Left Anthropometric Parameters of Ewes Stratified by Sex.

#### Pearson’s Correlation Between Height and Percutaneous Long Bone Lengths of Asante and Ewe Participants Stratified by Sex

Results of the Pearson’s correlation between height and percutaneous bone lengths of both ethnic groups based on sex are summarised in Tables 4 and 5.

**Table 4:** Pearson’s Correlation Between Height and Long Bone Lengths of Asantes Stratified by Sex.

|  | MALES (N = 71) |  | FEMALES (N = 69) |  |
| --- | --- | --- | --- | --- |
|  | HEIGHT |  | HEIGHT |  |
|  | r | p | r | p |
| RHL | 0.700 | 0.000 | 0.748 | 0.000 |
| RUL | 0.614 | 0.000 | 0.743 | 0.000 |
| RRL | 0.459 | 0.000 | 0.589 | 0.000 |
| RFL | 0.516 | 0.000 | 0.701 | 0.000 |
| RTL | 0.763 | 0.000 | 0.754 | 0.000 |
| RFibL | 0.829 | 0.000 | 0.785 | 0.000 |
| LHL | 0.703 | 0.000 | 0.741 | 0.000 |
| LUL | 0.645 | 0.000 | 0.813 | 0.000 |
| LRL | 0.479 | 0.005 | 0.562 | 0.000 |
| LFL | 0.504 | 0.000 | 0.654 | 0.000 |
| LTL | 0.822 | 0.000 | 0.727 | 0.000 |
| LFibL | 0.831 | 0.000 | 0.788 | 0.000 |

*RHL = Right Humeral Length; RUL = Right Ulnar Length; RRL = Right Radial Length; RFL = Right Femoral length; RTL = Right Tibial Length; RFibL = Right Fibular length; LHL = Left Humeral Length; LUL = Left Ulnar Length; LRL = Left Radial Length; LFL = Left Femoral length; LTL = Left Tibial length; LFibL = Left Fibular length; r = Pearson's correlation co-efficient; p = Statistical significance level*

**Table 5:** Pearson’s Correlation Between Height and Long Bone Lengths of Ewes Stratified by Sex.

|  | MALES (n=61) |  | FEMALES (n=41) |  |
| --- | --- | --- | --- | --- |
|  | HEIGHT |  | HEIGHT |  |
|  | r | p | r | p |
| RHL | 0.693 | 0.000 | 0.621 | 0.000 |
| RUL | 0.158 | 0.225 | 0.570 | 0.000 |
| RRL | 0.012 | 0.925 | 0.340 | 0.000 |
| RFL | 0.708 | 0.000 | 0.751 | 0.000 |
| RTL | 0.650 | 0.000 | 0.797 | 0.000 |
| RFibL | 0.753 | 0.000 | 0.759 | 0.000 |
| LHL | 0.702 | 0.000 | 0.588 | 0.000 |
| LUL | 0.117 | 0.370 | 0.513 | 0.000 |
| LRL | 0.040 | 0.762 | 0.329 | 0.036 |
| LFL | 0.723 | 0.000 | 0.724 | 0.000 |
| LTL | 0.693 | 0.000 | 0.723 | 0.000 |
| LFibL | 0.712 | 0.000 | 0.759 | 0.000 |

*RHL = Right Humeral Length; RUL = Right Ulnar Length; RRL = Right Radial Length; RFL = Right Femoral length; RTL = Right Tibial Length; RFibL = Right Fibular length; LHL = Left Humeral Length; LUL = Left Ulnar Length; LRL = Left Radial Length; LFL = Left Femoral length; LTL = Left Tibial length; LFibL = Left Fibular length; r = Pearson's correlation co-efficient; p = Statistical significance level*

### Linear regression analyses for height estimation using lengths of long bones

#### Regression Equations for Height Estimation Using Long Bone Lengths of Male Asantes and Ewes

Stepwise linear regression analysis was performed for height estimation for males using the percutaneous long bones of upper and lower limbs (Table 6).

**Table 6:**
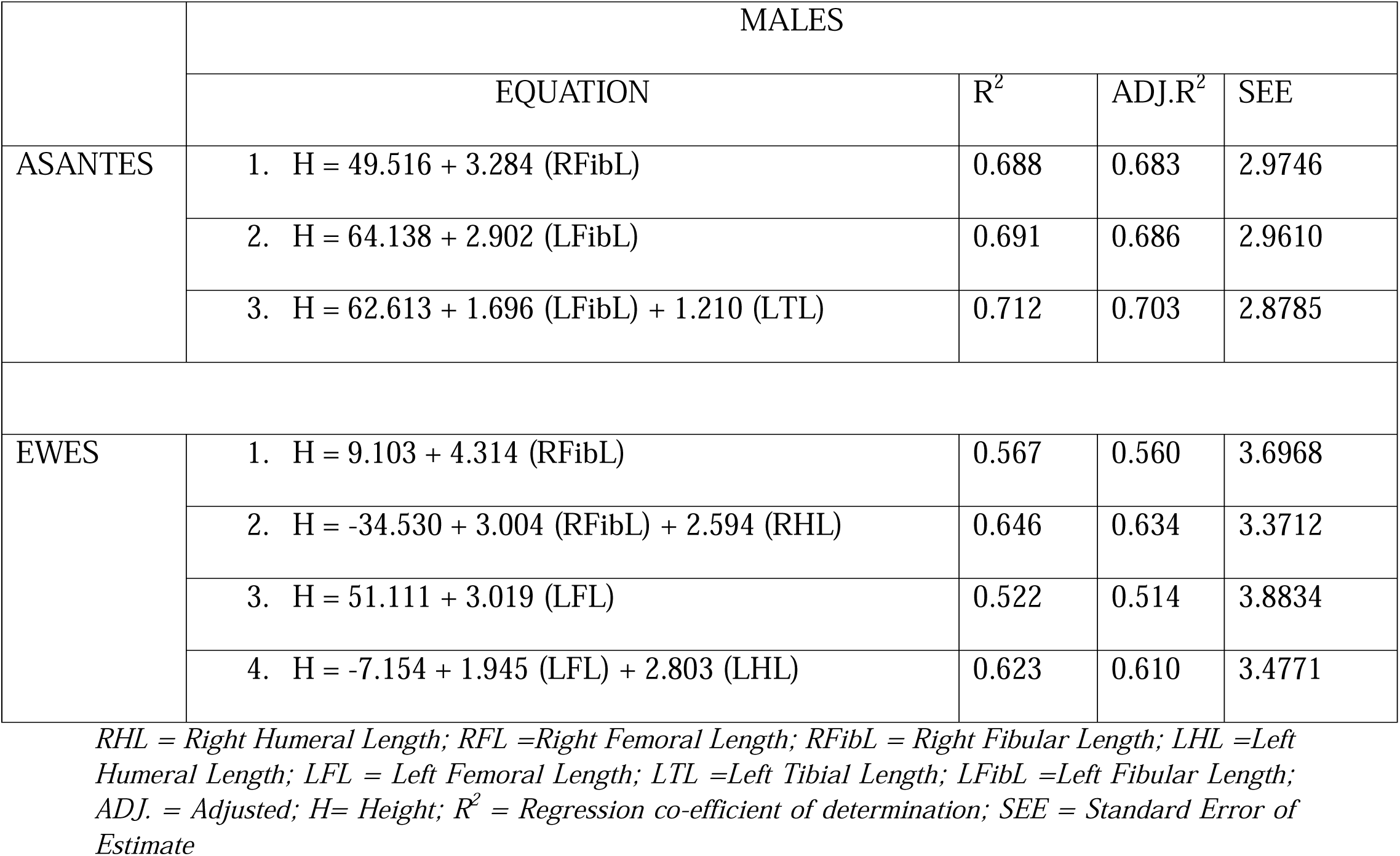
Linear Equations for Height Estimation Using Right Percutaneous Long Bone Lengths for The Males of Asantes and Ewes.

#### Regression Equations for Height Estimation Using Long Bone Lengths of Female Asantes and Ewes

Stepwise linear regression analysis was performed for height estimation for females using the lengths of percutaneous long bones of upper and lower limbs (Table 7).

**Table 7:** Linear Equations for Height Estimation Using Right Percutaneous Long Bone Lengths of Asantes and Ewes.

|  | FEMALES |  |  |  |
| --- | --- | --- | --- | --- |
|  | EQUATION | R<sup>2</sup> | ADJ.R<sup>2</sup> | SEE |
| ASANTES | 1. $H = 39.767 + 3.432 (\text{RFibL})$ | 0.616 | 0.610 | 4.1405 |
| | 2. $H = 32.041 + 2.303 (\text{RFibL}) + 1.778 (\text{RUL})$ | 0.722 | 0.714 | 3.5485 |
| | 3. $H = 70.706 + 3.338 (\text{LUL})$ | 0.660 | 0.655 | 3.8933 |
| | 4. $H = 42.532 + 2.090 (\text{LUL}) + 1.769 (\text{LFibL})$ | 0.742 | 0.734 | 3.4190 |
| | 5. $H = 43.658 + 2.495 (\text{LUL}) + 1.990 (\text{LFibL}) - 0.849 (\text{LRL})$ | 0.757 | 0.746 | 3.3404 |
| EWES | 1. $H = 26.521 + 3.585 (\text{RTL})$ | 0.636 | 0.626 | 2.6779 |
| | 2. $H = 25.980 + 2.954 (\text{RTL}) + 0.725 (\text{RHL})$ | 0.676 | 0.659 | 2.5587 |
| | 3. $H = 45.451 + 3.189 (\text{LFibL})$ | 0.576 | 0.565 | 2.8904 |

*RHL = Right Humeral Length; RUL = Right Ulnar Length; RFL = Right Femoral Length; RTL = Right Tibial Length; RFibL = Right Fibular Length; LUL = Left Ulnar Length; LRL = Left Radial Length; LFibL = Left Fibular Length; ADJ. = Adjusted; H= Height; R<sup>2</sup> = Regression co-efficient of determination; SEE = Standard Error of Estimate*

## Discussion

### Population characteristics of study participants

The present study comprised 140 Asantes [71(50.7%) males and 69(49.3%) females] and 102 Ewes [61(59.8%) males and 41(40.2%) females]. The age range of the participants of the study was 20 – 25 years. This age range was appropriate because studies have shown that, there is skeletal deformity with advancing age and thus reduction in one’s stature [15,16]. Also, it has been reported that, the ossification of bones of the limbs is completed around age 20 – 25 years of postnatal life. Other findings reveal that, beyond the fifth decade of extrauterine life, degenerative alterations occur in articulations and cartilages [17], consequently affecting limb dimensions and height. For instance, because participants aged above 65 years were recruited for height estimation using fibular length, unlike the present study which recorded strong correlation co-efficient between fibular length and height, that by Auyeung *et al.* [18] recorded moderate correlation among both males (r = 0.574) and females (r = 0.559).

In the present study, males were significantly taller than females among both Asantes and Ewes. This is in line with those reported by several investigators among different populations across the globe [19–23]. Similarly, Kavyashree *et al.* [24] reported significantly greater mean height among males (170.88 cm) than females (158.87 cm). Banerjee *et al.* [25] also had a significantly greater mean height for the male participants (164.05 cm) than the females (156.38 cm) which is in line with the present study. Although an individual’s stature is impacted significantly by epigenetic factors such as nutritional status, growth rate and physiological capacities, by and large, males are taller than females as a result of quite extended bone growth duration in the former than the latter [26]. This is perhaps due to the cessation of ossification of long bones of the limbs occurring earlier in females than in males [27]. Therefore, more precisely, males being taller than females could be directly linked to the early epiphyseal line formation in females as a result of high estradiol but low testosterone levels in females [28].

This is because, oestrogens are known for enhancing epiphyseal plate ageing process, causing the proliferative tendency of the plate chondrocytes to become exhausted leading to the synostotic epiphyseal line formation. As a result, males have about two more years of osseous tissue growth than females [20]. However, there was no statistically significant difference between Asante males and Ewe males or Asante females and Ewe females in terms of height (p > 0.05). This could be as a result of the closeness in stature which has been asserted by most Ghanaians although according to Chawla *et al.* [29], ethnic or race affiliation affects one’s height. This observation could be due to similar environmental conditions, common physical activities engaged in by study participants, genetic constitution and nutritional states which are essential in the growth and development of humans. All the participants of the present study were tertiary students residing in Kumasi as of the time of sampling. Being an inherent trait, stature forms one the most relevant identifying features of humans [30]. This is because, there are peculiarities in stature among different populations with respect to sex. Other studies have shown that stature of an individual is considerably influenced by age, sex, physical activity as well as the population [31,32].

### Sexual dimorphism of percutaneous bones of upper and lower limbs of the study participants

The present study recorded significantly greater mean percutaneous upper and lower limb bone lengths for both the right and left sides of the body among the males than females for both ethnic groups. These results also correspond to the differences in the heights of the study participants.

This is because, it has been asserted that, longer limbs are associated with taller people [33]. Regarding the ulnar length, the findings of the present study is in line with that by Paul *et al.* [17] who recruited 500 participants aged 20 – 50 years where males had a significantly longer ulnae than females although the upper limit of the age range differed significantly among the two studies. Another study by Itlapuram *et al.* [34] among the students of Narayana Medical College, Nellore, Andhra Pradesh, males recorded significantly longer ulnae than females. Regarding the humeral length, the present study is in conformity with findings of Ismail *et al.* [35] who reported sexual dimorphism in humeral lengths with males recording greater mean values than females among a Malaysian population. Also, in an Egyptian population, Ali and Abd Elbaky [36] reported significantly greater humeral length in males than females. Similarly, among South African Black indigenes, Whites and Hybrid populations, males recorded significantly greater values than females [37]. Males recording significantly greater mean humeral lengths than females could also be attributed to how often males are engaged in weeding and are involved in more physical activities using the upper limb than females at home and in school. More often, males actively engage in extracurricular activities such as playing football, volleyball, tennis among others. These activities could contribute to males having larger long bones than females. Additionally, in childhood, boys spend more time playing games that involve climbing, which may potentially extend their limbs and consequently lengthen their bones. Just like the current study, males had considerably longer mean femoral lengths than females [38]. It has been reported that, males have larger lower limbs compared to females, and this has been largely attributed to differences in environmental responses, mechanical loading patterns and hormonal differences [39]. To buttress this point, the literature has it that, the linear dimensions of especially the fibula in males are larger than that of females probably from the fact that, males have longer period of growth during puberty and therefore larger bones and musculature due to the influence of sex genes, sex steroid hormones and other hormones such as growth hormone and *insulin-like growth factor 1* alongside mechanical loading [40,41]. Moreover, it has been speculated that females have relatively shorter femora than males as a result of the former having broader pelves and thus greater bicondylar femoral angles [42]. In the present study, tibial lengths being greater in males than females are in line with reports from other studies [24,43]. For fibular length, findings of the present study are in line with that reported by Auyeung *et al.* [18] and Hishmat *et al.* [44] where males recorded significantly greater fibular lengths than females. However, although males recorded greater fibular length than females, no sexual dimorphism was observed among a Pakistani population [45].

### Estimation of height using percutaneous long bones of the limbs stratified by sex

In the present study, several differential regression models for height estimation for the Asante and Ewe participants were derived using lengths of long bones. Derivation of these formulae for various predictions is possible since there exist consistent biological relations between body parts and the entire body [46]. The most useful models for height estimation utilized fibular lengths. This was followed by ulnar lengths. This suggests that, based on head-to-head comparison, the lower limb percutaneous bones are cumulatively more useful than that of the upper limb. This contradicts a study carried out by Celbis and Agritmis [47] where it was opined that upper limb long bones are as useful as that of the lower limb for stature estimation of the living.

Where individuals have abnormal posture, their true height cannot be established. Therefore, the equations derived using the percutaneous long bones of both the upper and lower limbs can be applied conveniently for such applications. The equations are appropriate enough as both simple and multiple linear regression equations were derived for each group. This means that, the presence of only one of the long bones will not be a limitation to height estimation. According to Waghmare *et al.* [48], a particular long bone measurement or a combination of such bones is a reflection of the height of an individual. As with the present study, some authors have reported the appropriateness in the use of long bones for stature prediction since they are often associated with low standard error of estimates [49,50]. In the present study, the adjusted co-efficient of determinations of equations obtained for multivariate ones were greater than that of the univariate ones. This means that, multivariate equations are more useful than simple ones [46]. This is because, as evident in the present study, multiple regression shows reduced mean absolute percentage error of prediction [51].

According to Duyar and Pelin [52], for increased accuracy, regression equations should be population-specific. This is because, factors including ethnicity, occupation, nutrition and environment play significant roles in the overall development of humans. The staple food for Asantes is fufu (made using cassava) with varieties of soup while Ewes enjoy more of Akple (made using maize) with okra soap. In both food items, some pounding or riding using the upper limb is needed with concomitant support of the lower limbs. Environmental conditions affect the type of work members of a particular community embark on. Both Asantes and Ewes belong to the tropical area of Ghana and are mainly involved in agricultural activities. Therefore, there is some kind of closeness between the two ethnic groups with respect to their daily activities. However, for increased accuracy through elimination of other confounding factors, in the present study, models derived were sex-and ethnic-specific.

Although, it has been proposed that the best way to estimate height of an individual is to use skeletal remains, percutaneous measurements of bones in the living can also serve as surrogate means [2]. It has been shown that, stature estimation using bones is quite difficult and cumbersome since it involves several activities including cleaning and bone preparations. As a result, percutaneous measurements instead of direct bone measurements are often embraced and preferred by most forensic anthropologists since it is more advantageous [53]. That is why the present study considered the percutaneous approach just like other studies [54,55]. Reconstruction of an individual’s stature using body parts offers great anthropological merits when identifying even the dead. In addition, height estimation is useful during the stepwise identification of an unknown person since height is a useful feature essential to help bring down the window of search of individuals [56]. Congruent with other study findings, other authors have reported reliable results in the use of long bones for height estimation [57,58].

Among the population of Jordan and a study carried out in Northern Ghana, humeral length was found to be a proxy useful for height estimation [2,59]. Shah *et al.* [54] utilized regression method to derive models for the estimation of height using arm length, which is almost the same as percutaneous humeral length result of the present study. Additionally, the use of percutaneous humeral lengths or fragment measurements for height estimation has been reported by other investigators [36,60]. Stature predicted from measurements of body parts are very useful especially among individuals with disproportionate growth disorder, myopathy, skeletal dysplasia or stature loss during spinal column procedures [5]. This is because, it has been shown that, there is steady height increase of a person perhaps till adolescence level and then later declines due to backbone erosion occurring as one advances in age [17]. Therefore, in situations where some disease conditions or disabilities would not permit direct height measurement, the derived formulae would be suitable as proxy predictors of height. In the present study, among other parameters, right tibial length was useful for height estimation among Ewe females. This is in agreement with that by Kavyashree *et al.* [24]. Similarly, a study carried out on 400 participants aged 17-24 years, it was found out that, tibial length explained 74% of height variation in males and 72% in females [61]. In the present study, linear regression models were derived for each ethnic group and sex so as to satisfy the proven concept of interpopulation variation in body proportions [62]. Regression equations derived for the prediction of height in the present study did not differ significantly from the actual measured height of the study participants, making the derived models very appropriate for stature estimation.

## Conclusion

In both Asantes and Ewes, males were significantly taller and had greater anthropometric parameters than females. For the same sex, there was no statistically significant difference between Asantes and Ewes with respect to height. Ewe males had significantly greater ulnar and radial lengths than Asante males. Meanwhile, Ewe females had significantly greater ulnar, radial, femoral, tibial and fibular lengths than Asante females.

Most of the percutaneous long bones were useful for height estimation in the present study for both ethnic groups. The most useful parameter for the Asante males was left fibular and tibial lengths with that of the females being left ulnar, fibular and radial lengths. For the Ewe males, the most significant parameters for height estimation were right fibular and humeral lengths with the Ewe females being right tibial and humeral lengths.

The study has presented efficient anthropometric models for adequate identification of individuals by applying the appropriate corresponding model. Findings of the study are therefore useful for the biological profiling of humans with dismembered body parts involved in various disasters such as automobile accidents. The database and formulae derived would be useful for stature reconstruction needed for various assessments on bedridden patients.

## Acknowledgements

We express our profound appreciation to all persons who willingly took part in this study.

## Data Availability Statement

Data used to support the study findings would be made available by the corresponding author upon request.

## Author Contributions

**Conceptualization:** Daniel Kobina Okwan, Chrissie Stansie Abaidoo, Pet-Paul Wepeba, Juliet Robertson, Samuel Kwadwo Peprah Bempah

**Data curation:** Daniel Kobina Okwan, Priscilla Obeng, Ethel Akua Achiaa Domfeh, Sarah Owusu Afriyie, Thomas Kwaku Asante

**Methodology:** Daniel Kobina Okwan, Chrissie Stansie Abaidoo, Pet-Paul Wepeba, Juliet Robertson, Samuel Kwadwo Peprah Bempah, Priscilla Obeng, Ethel Akua Achiaa Domfeh

**Formal analysis:** Daniel Kobina Okwan, Chrissie Stansie Abaidoo, Pet-Paul Wepeba, Juliet Robertson, Samuel Kwadwo Peprah Bempah, Sarah Owusu Afriyie, Thomas Kwaku Asante

**Resources:** Daniel Kobina Okwan, Chrissie Stansie Abaidoo, Pet-Paul Wepeba, Thomas Kwaku Asante

**Validation:** Chrissie Stansie Abaidoo, Pet-Paul Wepeba, Daniel Kobina Okwan, Thomas Kwaku Asante

**Writing (original draft/editing/reviewing):** Daniel Kobina Okwan, Chrissie Stansie Abaidoo, Pet-Paul Wepeba, Juliet Robertson, Samuel Kwadwo Peprah Bempah, Priscilla Obeng, Ethel Akua Achiaa Domfeh, Sarah Owusu Afriyie, Thomas Kwaku Asante

## References

[1] Singh A, Nagar M, Kumar A. An anthropometric study of the humerus in adults. Research and Reviews: Journal of Medical and Health Sciences. 2014;3(3):77–82.

[2] Amidu N, Banyeh M, Bani SB, Adam Y, Dapare PP, Zogli KE. Models for predicting height from percutaneous lengths of the ulna and femur in a Ghanaian population. Canadian Society of Forensic Science Journal. 2021 Jan 2;54(1):49–60.

[3] Karadayı B, Kaya A, Koç H, Varlık E, Özaslan A. Sex determination for using hand and wrist measurements in Turkish population. Turkish Journal of Forensic Medicine. 2014;28(2):132–40.

[4] Ukoha UU, Umeasalugo KE, Udemezue OO, Asomugha LA. Estimation of stature from cephalic dimensions in a nigerian population. Estimación de la estatura por las dimensiones cefálicas en una población nigeriana. Revista Argentina de Anatomía Clínica. 2015;7(1):17–25.

[5] Navid S, Mokhtari T, Alizamir T, Arabkheradmand A, Hassanzadeh G. Determination of stature from upper arm length in medical students. Anatomical Sciences Journal. 2014 Aug 10;11(3):135–40.

[6] Mike IN, Ebe NM, Andrew OO, Ambrose OE. Percutaneous anthropometry of hand dimensions for stature reconstruction among Nigerians. Forensic Science and Addiction Research. 2018 Aug 6;3(5):1–8.

[7] Sládek V, Macháček J, Ruff CB, Schuplerová E, Přichystalová R, Hora M. Population□specific stature estimation from long bones in the early medieval Pohansko (Czech Republic). American journal of physical anthropology. 2015 Oct;158(2):312–24.

[8] Dumitru M, Mangaloiu D, Punga A, Lungu A. Factors affecting growth and bone development in minors. Case report: Madalina Dumitru. The European Journal of Public Health. 2015 Oct 1;25(suppl_3):ckv176-222.

[9] Patel SH, Bastia BK, Kumar L, Kumaran SM. Estimation of adult human stature from measurements of inter- acromial length in Gujarati population of India. Journal of Indian Academy of Forensic Medicine. 2015;37(4):365–8.

[10] Wankhede KP, Anjankar VP, Parchand MP, Kamdi NY, Patil ST. Estimation of stature from head length and head breadth in Central Indian population: an anthropometric study. Int J Anat Res. 2015 Mar 31;3(1):954–7.

[11] Sukumar CD. Estimation of stature based on percutaneous length of Ulna in living subjects. Sch J App Med Sci. 2017 May;5(5C):1897–902.

[12] Coleman A. Road traffic accidents in Ghana: a public health concern, and a call for action in Ghana,(and the sub-region). Open Journal of Preventive Medicine. 2014 Nov 13;4(11):822.

[13] Song-in K, Riengrojpitak S, Tiensuwan M. Estimation of stature from forearm length in Thai children. Journal of Food Health and Bioenvironmental Science. 2013;6(1):131–40.

[14] Ibegbu AO, David ET, Hamman WO, Umana UE, Musa SA. Hand length as a determinat of height in school children. Advances in life sciences. 2015;5(1):12–7.

[15] Arlappa N, Qureshi IA, Ravikumar BP, Balakrishna N, Qureshi MA. Arm span is an alternative to standing height for calculation of body mass index (BMI) amongst older adults. International Journal of Nutrition. 2016 Mar 28;2(1):12–24.

[16] Tella SH, Gallagher JC. Prevention and treatment of postmenopausal osteoporosis. The Journal of steroid biochemistry and molecular biology. 2014 Jul 1;142:155–70.

[17] Paul M, Sengupta O, Halder S, Panda UK. A study for estimation of human height from the length of ulna. Int J Res Rev. 2020;7(1):101–13.

[18] Auyeung TW, Lee JS, Kwok T, Leung JL, Leung PC, Woo J. Estimation of stature by measuring fibula and ulna bone length in 2443 older adults. The journal of nutrition, health & aging. 2009 Dec;13:931–6.

[19] Chavan SK, Chavan KD, Mumbre SS, Makhani CS. Stature and percutaneus tibial length: A correlational study in Maharashtrian population. Indian journal of forensic medicine and pathology. 2009 Jul;2(3):109–12.

[20] Okwan DK, Abaidoo CS, Tetteh J, Kusi A. Forensic application of hand and foot biometrics as models for height, weight and sex determination among Ghanaians. Int J Anat Res. 2020;8(2.3):7531-38.

[21] Osuna-Padilla IA, Borja-Magno AI, Leal-Escobar G, Verdugo-Hernandez S. Validation of predictive equations for weight and height using body circumferences In Mexican elderlys. Nutricion Hospitalaria. 2015 Dec 1;32(6):2898–902.

[22] Venkataraman R, Ranganathan L, Nirmal V, Kameshwaran J, Sheela CV, Renuka MV, Ramakrishnan N. Height measurement in the critically ill patient: A tall order in the critical care unit. Indian journal of critical care medicine: peer-reviewed, official publication of Indian Society of Critical Care Medicine. 2015 Nov;19(11):665.

[23] Villaverde□Gutiérrez C, Sánchez□López MJ, Ramirez□Rodrigo J, Ocaña□Peinado FM. Should arm span or height be used in calculating the BMI for the older people? Preliminary results. Journal of clinical nursing. 2015 Mar;24(5-6):817–23.

[24] Kavyashree AN, Bindurani MK, Asha KR, Lakshmi P. Estimation of stature by morphometry of percutaneous tibia. Indian Journal of Clinical Anatomy and Physiology. 2018 Jul;5(3):308–13.

[25] Banerjee M, Samanta C, Sangram S, Hota M, Kundu P, Mondal M, Ghosh R, Majumdar S. Estimation of human height from the length of tibia. Indian J Basic Appl Med Res. 2015 Dec;5(1):30–47.

[26] Datta Banik S. Arm span as a proxy measure for height and estimation of nutritional status: A study among Dhimals of Darjeeling in West Bengal India. Annals of human biology. 2011 Nov 1;38(6):728–35.

[27] Flinn EB, Strickland BK, Demarais S, Christiansen D. Age and gender affect epiphyseal closure in white-tailed deer. Southeastern Naturalist. 2013 Jun;12(2):297–306.

[28] Börjesson AE, Lagerquist MK, Windahl SH, Ohlsson C. The role of estrogen receptor α in the regulation of bone and growth plate cartilage. Cellular and Molecular Life Sciences. 2013 Nov;70:4023–37.

[29] Chawla S, Henshaw R, Seeger L, Choy E, Blay JY, Ferrari S, Kroep J, Grimer R, Reichardt P, Rutkowski P, Schuetze S. Safety and efficacy of denosumab for adults and skeletally mature adolescents with giant cell tumour of bone: interim analysis of an open-label, parallel-group, phase 2 study. The lancet oncology. 2013 Aug 1;14(9):901–8.

[30] Bhavna NS, Nath S. Use of lower limb measurements in reconstructing stature among Shia Muslims. Internet Journal of Biological Anthropology. 2009;2(2):86–97.

[31] Mondal MK, Jana TK, Giri S, Roy H. Height prediction from ulnar length in females: a study in Burdwan district of West Bengal (regression analysis). Journal of clinical and diagnostic research: JCDR. 2012 Oct;6(8):1401.

[32] Smith S, Okai I, Abaidoo CS, Acheampong E. Association of ABO blood group and body mass index: A cross□sectional study from a Ghanaian population. Journal of nutrition and metabolism. 2018;2018(1):8050152.

[33] Nandi ME, Olabiyi OA, Ibeabuchi NM, Okubike EA, Iheaza EC. Stature Reconstruction from Percutaneous Anthropometry of Long Bones of Upper Extremity of Nigerians in the University of Lagos. Arab Journal of Forensic Sciences & Forensic Medicine. 2018 Jun 7;1(7):869–80.

[34] Itlapuram MP, Jyothi C, Rajesham K, Subrahmanyam BV, Reddy G, Phanidra SV. Assessment of stature from the percutaneous measurement of ulna in healthy volunteers. Indian Journal of Forensic and Community Medicine. 2015;2(3):154–7.

[35] Ismail NA, Abd Khupur NH, Osman K, Mansar AH, Shafie MS, Mohd Nor F. Stature estimation in Malaysian population from radiographic measurements of upper limbs. Egyptian Journal of Forensic Sciences. 2018 Dec;8:1–5.

[36] Ali DM, Abd Elbaky FA. Sex identification and reconstruction of length of humerus from its fragments: an Egyptian study. Egyptian Journal of Forensic Sciences. 2016 Jun 1;6(2):48–55.

[37] Ndou R, Schepartz LA. Morphometric characteristics of the humerus and ulna in limbs bearing the supratrochlear aperture (STA). The Anatomical Record. 2016 Feb;299(2):220–33.

[38] Imai N, Funayama K, Suzuki H, Tsuchiya K, Nozaki A, Minato I, Miyasaka D, Endo N. Stature estimation formulae based on bony pelvic dimensions and femoral length. Homo. 2020 Apr 30;71(2):111–9.

[39] Pietrobelli A, Sorrentino R, Durante S, Marchi D, Benazzi S, Belcastro MG. Sexual dimorphism in the fibular extremities of Italians and South Africans of identified modern human skeletal collections: A geometric morphometric approach. Biology. 2022 Jul 19;11(7):1079.

[40] Callewaert F, Sinnesael M, Gielen E, Boonen S, Vanderschueren D. Skeletal sexual dimorphism: relative contribution of sex steroids, growth hormone-insulin-like growth factor-I (GH-IGF-I) and mechanical loading. J Endocrinol. 2010;207(2):127–34.

[41] Lopez-Costas O, Rissech C, Trancho G, Turbon D. Postnatal ontogenesis of the tibia. Implications for age and sex estimation. Forensic science international. 2012 Jan 10;214(1-3):207–e1.

[42] Sharma K, Mansur DI, Khanal K, Haque MK. Variation of carrying angle with age, sex, height and special reference to side. Kathmandu University Medical Journal. 2013;11(4):315-8.

[43] Sume BW. Estimation of body height from percutaneous length of tibia in Debre Markos University students, North West Ethiopia. Egyptian Journal of Forensic Sciences. 2019 Dec;9:1–8.

[44] Hishmat AM, Michiue T, Sogawa N, Oritani S, Ishikawa T, Fawzy IA, Hashem MA, Maeda H. Virtual CT morphometry of lower limb long bones for estimation of the sex and stature using postmortem Japanese adult data in forensic identification. International journal of legal medicine. 2015 Sep;129:1173–82.

[45] Zahid A, Shakir MA, Shireen R. Morphological and topographical anatomy of diaphyseal nutrient foramina of dried Pakistani fibulae. J Coll Physicians Surg Pak. 2015 Aug 1;25(8):560–3.

[46] Wube B, Seyoum G, Taye G. Estimation of stature by anatomical anthropometric parameters in first-year regular undergraduate students at Debre Markos University, North West Ethiopia. Ethiopian Journal of Health Development. 2019;33(3).

[47] Celbis O, Agritmis H. Estimation of stature and determination of sex from radial and ulnar bone lengths in a Turkish corpse sample. Forensic Science International. 2006 May 10;158(2-3):135–9.

[48] Waghmare V, Gaikwad R, Herekar N. Estimation of the stature from the anthropometric measurement of hand length. The internet journal of biological anthropology. 2011;4(2):167–72.

[49] Mahakizadeh S, Moghani-Ghoroghi F, Moshkdanian G, Mokhtari T, Hassanzadeh G. The determination of correlation between stature and upper limb and hand measurements in Iranian adults. Forensic science international. 2016 Mar 1;260:27–30.

[50] Pal A, De S, Sengupta P, Maity P, Dhara PC. Estimation of stature from hand dimensions in Bengalee population, West Bengal, India. Egyptian Journal of Forensic Sciences. 2016 Jun 1;6(2):90–8.

[51] Rani D, Krishan K, Kumar A, Kanchan T. Assessment of body weight from percutaneous widths of the bones and joints-Implications in forensic and clinical examinations. Acta Bio Medica: Atenei Parmensis. 2021;92(3).

[52] Duyar İ, Pelin C. Estimating body height from ulna length: need of a population-specific formula. Eurasian journal of Anthropology. 2010 Feb 13;1(1):11–7.

[53] Amit K, Srivastava AK, Verma AK. Estimation of stature by percutaneous measurements of distal half of upper limb (forearm & hand). J Indian Acad Forensic Med. 2010;32(4):325–8.

[54] Shah T, Patel M, Nath S, Menon SK. A model for construction of height and sex from shoulder width, arm length and foot length by regression method. Journal of Forensic Science & Criminology. 2015 Feb 27;3(1):102.

[55] Sheikhazadi A, Hassanzadeh G, Mokhtari T, Sheikhazadi E, Saberi Anary SH, Qoreishy M. Stature estimation from percutaneous Tibia height: study of Iranian medical students. Joint and Bone Science Journal. 2015;2(2):121–7.

[56] Singh I, Varte LR, Rawat S. Estimation of stature from different anthropometric measurements of Gorkha soldiers. Defence Life Science Journal. 2019;4(1):33–7.

[57] Chandra A, Chandna P, Deswal S, Mishra RK, Kumar R. Stature prediction model based on hand anthropometry. International Journal of Biomedical and Biological Engineering. 2015 Apr 2;9(2):201–7.

[58] Mehta AA, Mehta AA, Gajbhiye VM, Verma S. Correlation of Percutaneous Tibial Length with Body Height and Estimation of Stature in Living Central India Population.

[59] Mashali AA, Taweel OE, Ekladious E. Stature prediction from anthropometry of extremities among Jordanians. J. Forensic Sci. Criminol. 2017;5(2):202.

[60] Salles AD, Carvalho CR, Silva DM, Santana LA. Reconstruction of humeral length from measurements of its proximal and distal fragments. Journal of Morphological Sciences. 2017 Jan 16;26(2):0-.

[61] Khatun SS, Sharma N, Jain SK, Gupta A. Estimation of stature from percutaneous tibial length in Indian population. Int J Anat Res. 2016;4(3):2571–6.

[62] Carretero JM, Rodríguez L, García-González R, Arsuaga JL, Gómez-Olivencia A, Lorenzo C, Bonmatí A, Gracia A, Martínez I, Quam R. Stature estimation from complete long bones in the Middle Pleistocene humans from the Sima de los Huesos, Sierra de Atapuerca (Spain). Journal of Human Evolution. 2012 Feb 1;62(2):242–55.

